# The impact of health literacy-sensitive design and heart age in a cardiovascular disease prevention decision aid: randomised controlled trial and end user testing

**DOI:** 10.1101/2021.09.20.21263868

**Authors:** Carissa Bonner, Carys Batcup, Julie Ayre, Erin Cvejic, Lyndal Trevena, Kirsten McCaffery, Jenny Doust

**Author notes:** **Correspondence:** Dr Carissa Bonner, Rm 128A, Edward Ford Building A27 | The University of Sydney | NSW | 2006, |, E. **Ethics approval:** This study has ethics approval from the University of Sydney Human Research Ethics Committee (ID 2019/774). **Trial registration:** The trial protocol was pre-registered at ANZCTR (Trial number ACTRN12620000806965).

## Abstract

**Introduction:** Shared decision making is as an essential principle for cardiovascular disease (CVD) prevention, where asymptomatic people are considering lifelong medication and lifestyle changes. This project aimed to develop and evaluate the first literacy-sensitive CVD prevention decision aid (DA) developed for people with low health literacy, and investigate the impact of literacy-sensitive design and heart age.

**Methods:** We developed the standard DA based on international standards. The literacy-sensitive version included simple language, supporting images, white space and a lifestyle action plan. A randomised trial included 859 people aged 45-74 using a 3 (DA: standard, literacy-sensitive, control) x 2 (heart age: heart age + percentage risk, percentage risk only) factorial design, with outcomes including prevention intentions/behaviours, gist/verbatim knowledge of risk, credibility, emotional response and decisional conflict. We iteratively improved the literacy-sensitive version based on end user testing interviews with 20 people with varying health literacy levels.

**Results:** Immediately post-intervention (n=859), there were no differences between the DA groups on any outcome. The heart age group was less likely to have a positive emotional response, perceived the message as less credible, and had higher gist/verbatim knowledge of heart age risk but not percentage risk. After 4 weeks (n=596), the DA groups had better gist knowledge of percentage risk than control. The literacy-sensitive decision aid group had higher fruit consumption, and the standard decision aid group had better verbatim knowledge of percentage risk. Verbatim knowledge was higher for heart age than percentage risk amongst those who received both.

**Discussion:** The literacy-sensitive DA resulted in increased knowledge and lifestyle change for participants with varying health literacy levels and CVD risk results. Adding heart age did not increase lifestyle change intentions or behaviour but did affect psychological outcomes, consistent with previous findings.

**MeSH Terms:** Health Literacy, Cardiovascular Diseases, Decision Making (Shared), Life Style, Decision Support Techniques

## INTRODUCTION

Prevention of cardiovascular disease (CVD) includes lifestyle interventions and medication for those at highest risk who are most likely to benefit. An “absolute risk” approach is supported by clinical evidence and endorsed by most national guidelines around the world^1–5^. The absolute risk of a heart attack or stroke in the next 5-10 years can be assessed using widely available calculators^1^, but there is substantial underuse of these tools in practice^6–11^. Providing medication to high risk and not low risk patients is a cost-effective approach^6^. However, up to 75% of high risk patients are not receiving recommended medication to prevent death and disability from CVD, while 25% of low risk patients are taking medication they are very unlikely to benefit from^7^. Recent guideline changes have led to calls for a shared decision making approach, to ensure medication prescribing for blood pressure and cholesterol is more in line with patient values^12–14^.

We also know that health literacy plays a role in CVD prevention. Low health literacy is common in many countries, with estimates ranging from 36 – 60% of the population in Australia, Europe and the US^15–17^. This is associated with poorer self-management, less access to the health system, increased chronic disease including CVD, and increased mortality^18^. It is therefore important to specifically engage this group in communication strategies about CVD prevention. This requires changes to the design of online patient resources, since most consumers seek health information online, but fewer than 1% of health information websites meet recommended readability levels; Grade 8 is recommended to meet the needs of people with varying health literacy^19,20^.

Some countries have used online CVD risk assessment tools for absolute risk and heart age to engage consumers in CVD prevention, with millions of users worldwide^21–24^. However, our systematic review of 73 online CVD risk assessment tools available to consumers found they were not suitable for people with lower health literacy: their readability level was too high; they frequently used unexplained medical terms; few used best practice risk communication formats such as frequencies in icon arrays; and they rated poorly on actionability (i.e. clarity in instructions of what actions/steps to take), which makes it hard for the average person to know what to do about the risk assessment result^25^. Our review of 25 online decision aids for CVD prevention found similar issues with understandability and actionability^26^, and few included lifestyle change as an option reduce risk with many focusing on medication only.

There are several evidence-based strategies to address the issue of communicating CVD risk to people with lower health literacy:

1. Use literacy-sensitive design to improve the readability of health information and reduce the cognitive load of action plans for behaviour change^27–29^.
2. Use best practice risk communication formats to explain abstract probabilities (e.g. 16%) using icon arrays and more concrete frequencies (e.g. 16 out of 100 people like you)^30–33^.
3. Use patient decision aids to improve understanding and decision making, including both lifestyle change and medication as clear actions that patients can take to reduce their risk^27,34,35^.

This project aimed to develop and test a new consumer engagement tool for CVD prevention based on the above strategies, to address the needs of Australians with different levels of health literacy. It builds on our previous development of a GP-focused risk calculator and decision aid^36^, and evaluation of the national heart age calculator^24^.

## METHODS

### Stage 1: Develop consumer engagement tool

In stage one we developed the literacy-sensitive version of our existing GP decision aid^37^, which calculates 5 year risk of a CVD event based on current guidelines^1^ and shows the effects of 9 lifestyle, medication and supplement interventions^36^. We added heart age to the absolute CVD risk calculation based on published methods^38^. The literacy-sensitive design included simple language, supporting images and white space to improve readability and understandability^28^, and a novel action plan format our team developed that has been shown to reduce unhealthy lifestyle behaviours amongst people with low health literacy^29^. We added options for physical activity and smoking to the existing tool to reduce unhealthy snacking, drawing on previous literature on effective if-then plans in these areas^39,40^.

### Stage 2: Randomised trial to identify best formats for low vs high health literacy

#### Design

The randomised trial was based on a 3×2 factorial design to test the effect of literacy-sensitive design (literacy-sensitive DA, standard DA, or control: Heart Foundation patient information) and risk format (explaining CVD risk only (as a percentage risk), or CVD risk percentage + heart age) on psychological and behavioural outcomes. See Table 1/Figure 1 for study design, and Figure 2/Appendix 1 for example intervention content. The trial was pre-registered at ANZCTR (Trial number ACTRN12620000806965).

**Table 1:** The 2×3 study design.

| **Group** | **Risk results** | **Decision aid** | **Action plan** |
| --- | --- | --- | --- |
| **Control HF information**<br>**risk % +**<br>**heart age** | Absolute percentage risk shown in the style of HF risk calculator results<sup>41</sup>.<br>Heart age also shown in style of HF heart age calculator. | In the style of the NVDPA risk calculator<sup>41</sup>: participants can change any risk factors, then are presented with their risk percentage compared with their 'updated risk' based on the changes they made to risk factors. They are then advised to book in for a heart health check with their doctor. | Participants receive feedback on their blood pressure, cholesterol and BMI. Then they are prompted to select a topic to see more information about (diet, exercise or smoking). This was from the HF website<sup>42-44</sup>. |
| **Control HF information**<br>**risk % only** | Same as above but with no heart age. |  |  |
| **Standard DA**<br>**risk % +**<br>**heart age** | Absolute percentage risk shown alongside an icon array, with the number of icons in red (out of 100 grey icons) demonstrating the risk percentage.<br>Heart age also shown in style of HF heart age calculator. | Participants were asked to choose an option to reduce their risk, out of 9 potential options in 3 categories (medication, lifestyle changes and supplements). Once they chose an option, they were shown an icon array with the new risk in red, and the difference between their current and new risk in green. They were then shown information from our current CVD risk website about the option they chose, as well as a table of | Participants had to choose a lifestyle behaviour change to make (smoking, exercise or diet) and then create a goal. They were then guided through creating a 'SMART goal' style plan, taken from our current CVD risk website<sup>37</sup>. |
| **Standard DA**<br>**risk % only** | Same as above but with no heart age. |  |  |
|  |  | the benefits and harms of that choice<sup>37</sup>. |  |
| **Literacy-sensitive DA risk % + heart age** | Absolute percentage risk shown alongside an icon array, with the number of icons in red (out of 100 grey icons) demonstrating the risk percentage.<br>Heart age also shown with more explanation than control and standard DA conditions. | The same as for the standard decision aid above, however the information and benefits and harms were edited to be appropriate for all levels of health literacy, for example by introducing white space, images, and reducing the readability level. | Participants were prompted to change either their smoking, exercise or snacking habits. They were then guided through creating an action plan based on implementation intentions, or 'if-then' plans. The snacking action plan was previously developed by our team<sup>29</sup>, and the exercise and smoking plans were in the same style using research in those areas<sup>39,40</sup>. |
| **Literacy-sensitive DA risk % only** | Same as above but with no heart age. |  |  |

DA: decision aid, HF: Heart Foundation

**Figure 1:**
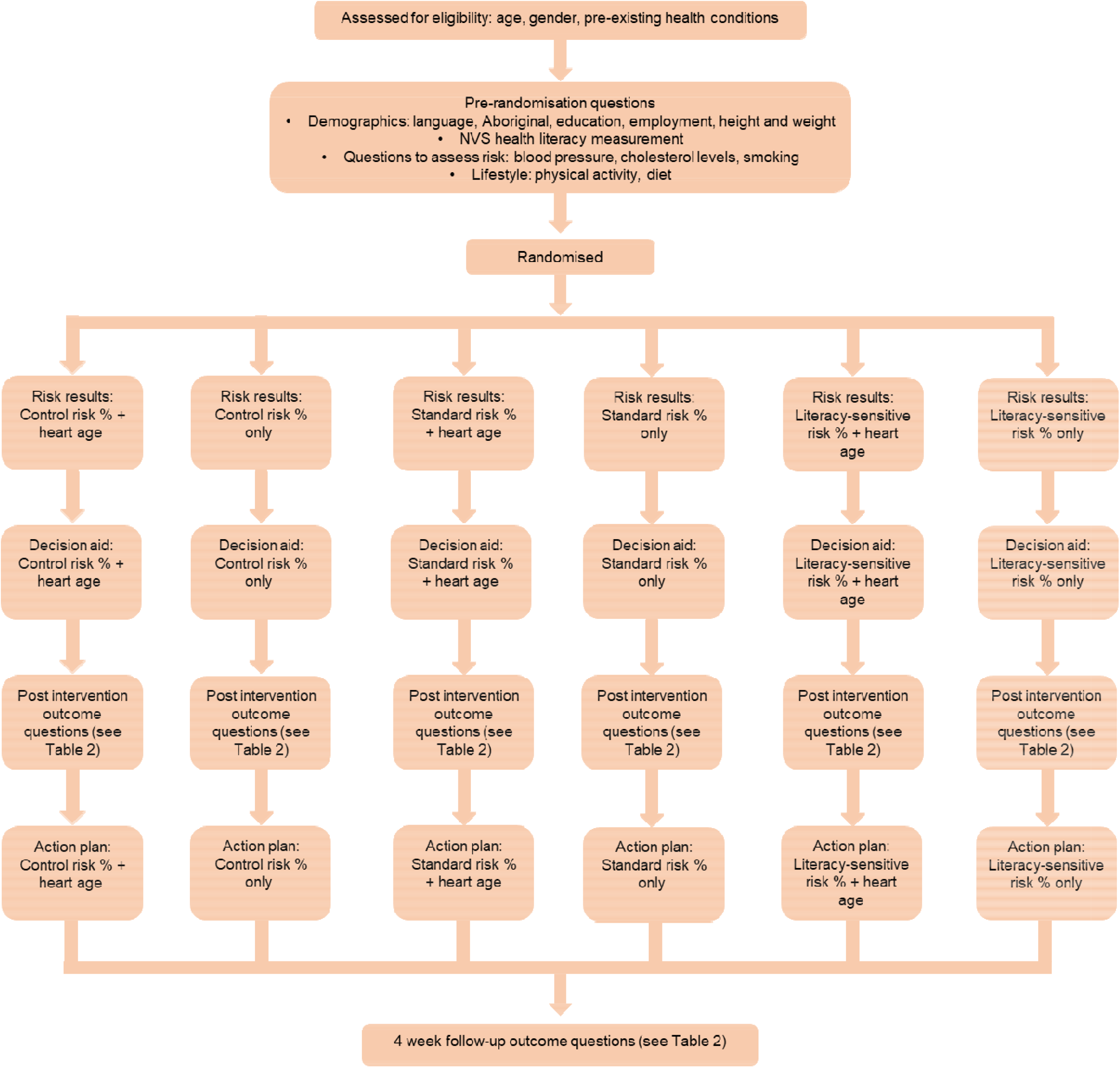
Study design.

**Figure 2:**
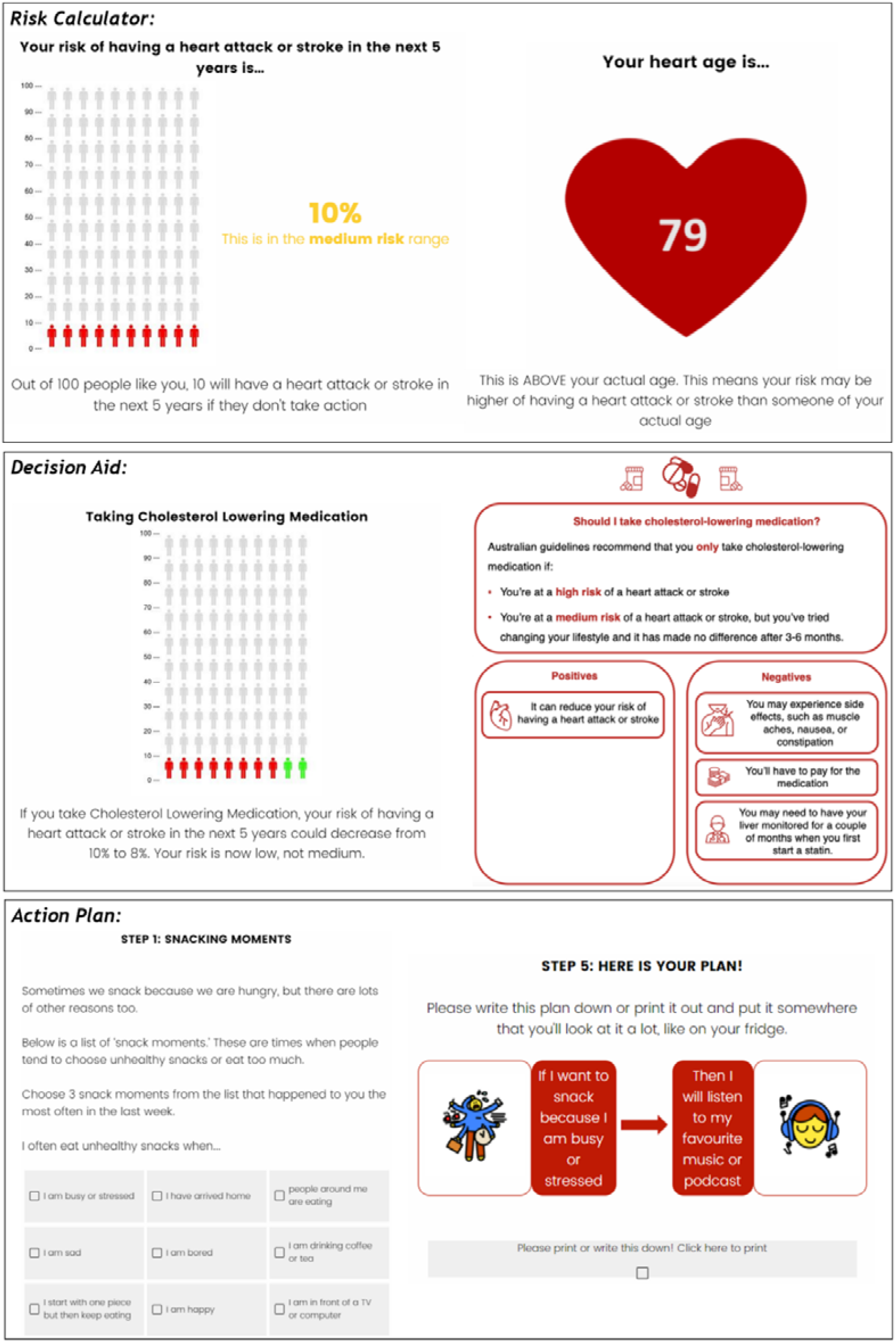
Example risk calculator, decision aid & action plan (literacy-sensitive heart age version)

#### Recruitment

A national sample was recruited through Qualtrics, an online social research agency, with stratified sampling based on gender/age groups (5 year age groups from 45-74 years). Participants completed a CVD risk assessment based on Australian guidelines and the New Zealand approach to calculating heart age^1,38^. If blood pressure or cholesterol were not known, the average by age/gender based on non-diabetic participants in the AusDiab cohort was used (accessed via JD), and all participants were advised to see a GP for a more accurate risk assessment. Those with established CVD or taking CVD prevention medication were excluded. Duplicate IP addresses were replaced, and stratified sampling was relaxed with additional quality checks added if hard to reach groups did not reach quota after 2 weeks.

#### Measures

Established measures were used for the primary outcome of behavioural intentions (validated Theory of Planned Behaviour scale applied to smoking, diet, exercise and GP visit)^45–47^. Secondary outcomes included: self-reported behaviour after 4 weeks compared to national guidelines for diet and physical activity^46,47^; gist and verbatim knowledge (absolute risk percentage and heart age); emotional response using the PANAS scale (three positive emotions e.g. hopeful, three negative emotions e.g. anxious)^48^; credibility of the information (that the information is personally relevant)^49^; and decision conflict scale (uncertainty in decision making)^50^. Full details are in Table 2.

**Table 2:**
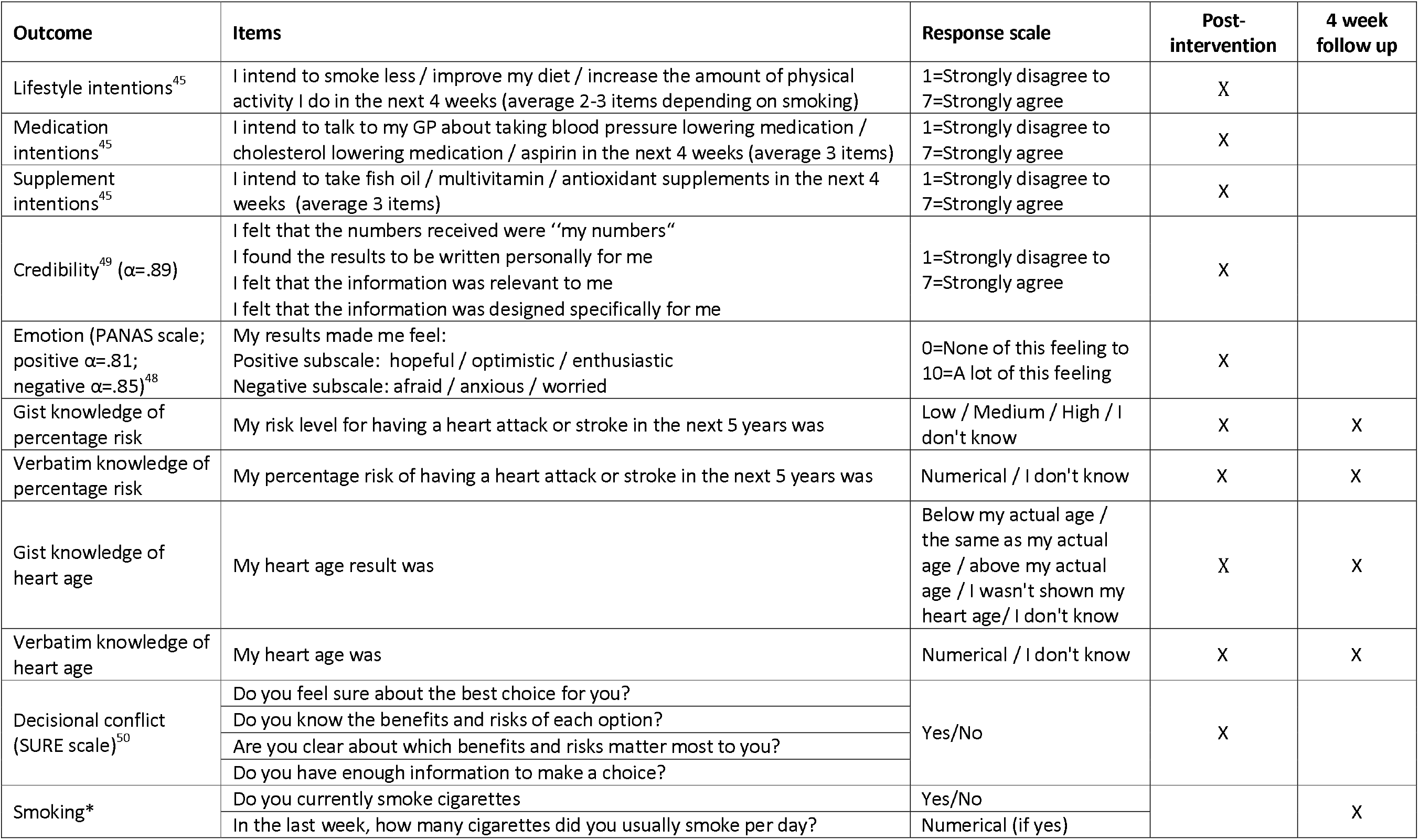
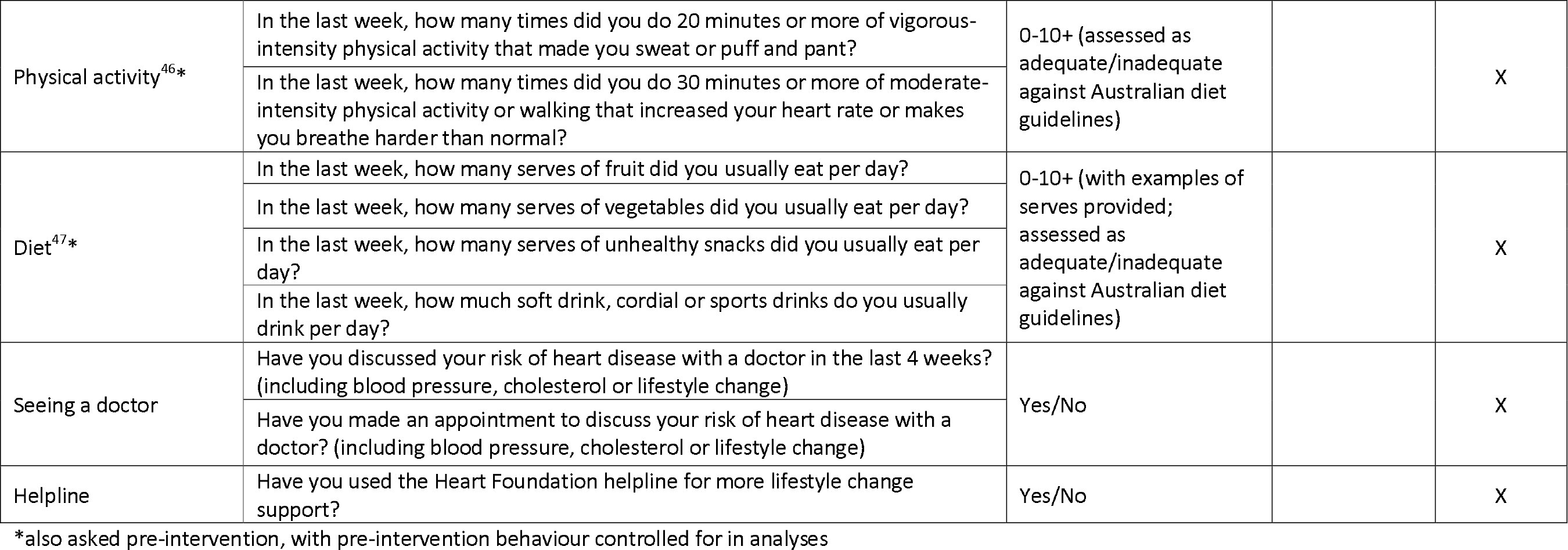
Psychological and behavioural outcomes measured in the analyses.

| Outcome | Items | Response scale | Post-intervention | 4 week follow up |
| --- | --- | --- | --- | --- |
| Lifestyle intentions<sup>45</sup> | I intend to smoke less / improve my diet / increase the amount of physical activity I do in the next 4 weeks (average 2-3 items depending on smoking) | 1=Strongly disagree to 7=Strongly agree | X |  |
| Medication intentions<sup>45</sup> | I intend to talk to my GP about taking blood pressure lowering medication / cholesterol lowering medication / aspirin in the next 4 weeks (average 3 items) | 1=Strongly disagree to 7=Strongly agree | X |  |
| Supplement intentions<sup>45</sup> | I intend to take fish oil / multivitamin / antioxidant supplements in the next 4 weeks (average 3 items) | 1=Strongly disagree to 7=Strongly agree | X |  |
| Credibility<sup>49</sup> ($\alpha=.89$) | I felt that the numbers received were “my numbers”<br>I found the results to be written personally for me<br>I felt that the information was relevant to me<br>I felt that the information was designed specifically for me | 1=Strongly disagree to 7=Strongly agree | X | |
| Emotion (PANAS scale; positive $\alpha=.81$; negative $\alpha=.85$)<sup>48</sup> | My results made me feel:<br>Positive subscale: hopeful / optimistic / enthusiastic<br>Negative subscale: afraid / anxious / worried | 0=None of this feeling to 10=A lot of this feeling | X | |
| Gist knowledge of percentage risk | My risk level for having a heart attack or stroke in the next 5 years was | Low / Medium / High / I don't know | X | X |
| Verbatim knowledge of percentage risk | My percentage risk of having a heart attack or stroke in the next 5 years was | Numerical / I don't know | X | X |
| Gist knowledge of heart age | My heart age result was | Below my actual age / the same as my actual age / above my actual age / I wasn't shown my heart age / I don't know | X | X |
| Verbatim knowledge of heart age | My heart age was | Numerical / I don't know | X | X |
| Decisional conflict (SURE scale)<sup>50</sup> | Do you feel sure about the best choice for you? | Yes/No | X |  |
|  | Do you know the benefits and risks of each option? |  |  |  |
|  | Are you clear about which benefits and risks matter most to you? |  |  |  |
|  | Do you have enough information to make a choice? |  |  |  |
| Smoking* | Do you currently smoke cigarettes | Yes/No |  | X |
|  | In the last week, how many cigarettes did you usually smoke per day? | Numerical (if yes) |  |  |
| Physical activity<sup>46*</sup> | In the last week, how many times did you do 20 minutes or more of vigorous-intensity physical activity that made you sweat or puff and pant? | 0-10+ (assessed as adequate/inadequate against Australian diet guidelines) |  | X |
|  | In the last week, how many times did you do 30 minutes or more of moderate-intensity physical activity or walking that increased your heart rate or makes you breathe harder than normal? |  |  |  |
| Diet<sup>47*</sup> | In the last week, how many serves of fruit did you usually eat per day? | 0-10+ (with examples of serves provided; assessed as adequate/inadequate against Australian diet guidelines) |  | X |
|  | In the last week, how many serves of vegetables did you usually eat per day? |  |  |  |
|  | In the last week, how many serves of unhealthy snacks did you usually eat per day? |  |  |  |
|  | In the last week, how much soft drink, cordial or sports drinks do you usually drink per day? |  |  |  |
| Seeing a doctor | Have you discussed your risk of heart disease with a doctor in the last 4 weeks? (including blood pressure, cholesterol or lifestyle change) | Yes/No |  | X |
|  | Have you made an appointment to discuss your risk of heart disease with a doctor? (including blood pressure, cholesterol or lifestyle change) |  |  |  |
| Helpline | Have you used the Heart Foundation helpline for more lifestyle change support? | Yes/No |  | X |

\*also asked pre-intervention, with pre-intervention behaviour controlled for in analyses

#### Analysis

A priori sample size calculations determined that 85 participants per randomised group (total n=510) would yield 90% power to detect a moderate effect size of d=0.5 (standardised difference) in the primary outcome of intention to change lifestyle or any of the secondary outcomes, assuming a two-sided alpha of 0.05. We aimed to recruit 20% more cases to account for potential missing values, totalling 600 participants (100 per group) at follow-up. This sample was inflated for recruitment to 850 to account for potential attrition of up to 30% between the intervention and follow-up.

Continuous outcome variables were modelled using linear regression. Dichotomous outcomes were analysed using modified Poisson regression (using a log-link function with robust error variances). Ordinal logistic regression was used to analyse ordered categorical outcomes. Count outcomes were modelled using negative binomial regression. All regression models included decision aid group (literacy-sensitive decision aid; standard decision aid; or basic Heart Foundation patient information) and risk format (CVD risk percentage only, or CVD risk percentage + heart age) as categorical variables and controlled for health literacy adequacy (categorical based on the Newest Vital Sign (NVS) measure^51,52^: low; moderate; adequate) and absolute risk (percentage). Post-intervention and follow-up outcomes were analysed separately, with follow-up analyses controlling for pre-intervention values where available. Pairwise comparisons were conducted to address the hypotheses. We also conducted exploratory analyses of potential differences in decision aid effects between health literacy levels by including a literacy-sensitive-by-decision aid interaction term, and by heart age category for heart age groups (younger/same vs older in stratified analyses). McNemar’s Chi-squared test for paired proportions was used to compare knowledge of heart age versus percentage risk amongst those who saw both. Analyses were conducted using Stata/IC v16.1 (StataCorp, College Station, TX, USA).

#### Hypotheses

1. The 2 decision aid formats will be more effective than standard Heart Foundation information;
2. The literacy-sensitive decision aid will be more effective than the standard decision aid for everyone (not just people with lower health literacy);
3. Adding heart age to absolute risk will be more effective than absolute risk alone.

### Stage 3: Iterative end user testing with varying health literacy levels

Participants in the trial were invited to opt-in to a “think aloud” interview to provide further end user testing and feedback for the literacy-sensitive version of the intervention. Participants went through the risk calculator in full, while saying out loud everything they were thinking, for example any areas of confusion. Further questions were also asked to prompt more discussion or elaboration. Transcripts were thematically coded and discussed after each set of 4-5 interviews, and improvements were made to the intervention before the next set of interviews. We conducted 2 rounds of interviews for people with low health literacy as our key target group (n=8), and then tested the improved tool with people who had higher health literacy to ensure it was suitable for these users in another two rounds (n=12).

## RESULTS

### Stage 1

We used the question format and style of the current national heart age calculator as the basis for the risk factor questions in all groups, and also based the heart age presentation on that tool. The CVD risk results and decision aid were presented based on: 1) our existing GP decision aid tool^37^ (standard decision aid group); 2) a simplified version of the standard decision aid with supporting images (literacy-sensitive decision aid group, see Figure 2); and 3) the current risk calculator from the National Vascular Disease Prevention Alliance^41^. See Appendix 1 for example intervention content in each group.

### Stage 2

The CONSORT diagram is shown in Figure 3, and characteristics of all participant groups in the intervention are shown in Table 3. We conducted a soft launch of n=100 participants to check we had an adequate low health literacy sample and adequate follow-up considering COVID-19 disruptions in 2020, before proceeding with the full trial with no changes to the pre-registered method. We recruited 859 participants for the intervention (including the 100 in the soft launch), with a target of 600 at 4 week follow-up, for which we recruited 596. Characteristics were similar between groups for age and gender, but some differences were observed for health literacy (relating to education) and absolute risk (relating to smoking and heart age), so these two factors were controlled for in the analyses. In terms of drop-out, there was no difference in randomised decision aid group (p=.71), randomised to heart age shown (p=.91), health literacy level (p=.69), CVD risk level (p=.56), or heart age result (p=.30) between those who returned for follow-up and those who did not. Outcomes by trial group are shown in Table 4; and analyses for each of the 3 hypotheses are shown in Tables 5-7.

**Table 3:** Trial participant characteristics by randomised group.

| Characteristics | Decision aid (DA) group |  |  | Heart age group |  |
| --- | --- | --- | --- | --- | --- |
|  | Control<br>(n=290) | Standard<br>(n=285) | Literacy-sensitive<br>(n=284) | Risk % only<br>(n=432) | Risk % +<br>heart age<br>(n=427) |
| **Demographics** |  |  |  |  |  |
| Age (years), mean (SD) | 59.9 (8.7) | 59.6 (8.2) | 58.3 (8.7) | 58.8 (8.6) | 59.8 (8.5) |
| Heart age (years), mean (SD) | 60.7 (14.7) | 60.9 (13.1) | 58.5 (13.7) | 58.9 (14.0) | 61.2 (13.7) |
| Male, n (%) | 137 (47.2%) | 147 (51.6%) | 142 (50.0%) | 213 (49.3%) | 213 (49.9%) |
| Female, n (%) | 153 (52.8%) | 138 (48.4%) | 142 (50.0%) | 219 (50.7%) | 214 (50.1%) |
| University degree, n (%) | 149 (51.4%) | 133 (46.7%) | 145 (51.1%) | 218 (50.5%) | 209 (48.9%) |
| Inadequate health literacy, n (%) | 63 (21.7%) | 77 (27.0%) | 66 (23.2%) | 103 (23.8%) | 103 (24.1%) |
| **Clinical characteristics** |  |  |  |  |  |
| Knew their cholesterol, n (%) | 41 (14.1%) | 41 (14.4%) | 34 (12.0%) | 59 (13.7%) | 57 (13.3%) |
| Total cholesterol<sup>^</sup>, mean (SD) | 4.9 (1.3) | 4.9 (1.5) | 4.4 (1.4) | 4.6 (1.3) | 4.8 (1.5) |
| HDL cholesterol<sup>^</sup>, mean (SD) | 2.6 (1.3) | 2.6 (1.2) | 2.8 (1.3) | 2.8 (1.3) | 2.6 (1.2) |
| Knew their blood pressure, n (%) | 102 (35.2%) | 106 (37.2%) | 98 (34.5%) | 162 (37.5%) | 144 (33.7%) |
| Systolic blood pressure<sup>^</sup>, mean (SD) | 123.9 (15.1) | 127.0 (14.8) | 124.9 (14.8) | 123.9 (14.7) | 126.8 (15.1) |
| Diastolic blood pressure<sup>^</sup>, mean (SD) | 83.1 (11.7) | 83.3 (12.0) | 82.3 (13.0) | 82.4 (12.7) | 83.5 (11.8) |
| Overweight BMI<sup>*</sup>, n (%) | 172 (59.3%) | 175 (61.4%) | 161 (56.7%) | 260 (60.2%) | 248 (58.1%) |
| **Behaviour + lifestyle characteristics** |  |  |  |  |  |
| Adequate diet<sup>*</sup>, n (%) | 73 (25.2%) | 75 (26.3%) | 67 (23.6%) | 113 (26.2%) | 102 (23.9%) |
| Adequate exercise<sup>*</sup>, n (%) | 165 (56.9%) | 150 (52.6%) | 162 (57.0%) | 239 (55.3%) | 238 (55.7%) |
| Smokers, n (%) | 38 (13.1%) | 42 (14.7%) | 35 (12.3%) | 48 (11.1%) | 67 (15.7%) |
| **Risk results** |  |  |  |  |  |
| Older heart age<sup>#</sup>, n (%) | 164 (56.6%) | 171 (60.0%) | 153 (53.9%) | 230 (53.2%) | 258 (60.4%) |
| Absolute risk, mean (SD) | 5.3 (4.8) | 5.4 (4.1) | 4.9 (4.1) | 4.9 (4.5) | 5.5 (4.2) |
| Low risk, n (%) | 248 (85.5%) | 235 (82.5%) | 238 (83.8%) | 375 (86.8%) | 346 (81.0%) |
| Medium risk, n (%) | 37 (12.8%) | 44 (15.4%) | 39 (13.7%) | 49 (11.3%) | 71 (16.6%) |
| High risk, n (%) | 5 (1.7%) | 6 (2.1%) | 7 (2.5%) | 8 (1.9%) | 10 (2.3%) |

<sup>\*</sup> overweight BMI > 25; adequate diet = at least 2 servings of fruit and 5 servings of vegetables per day in the past week<sup>47</sup>; adequate physical activity = 3 vigorous sessions per week, 5 moderate sessions per week, or 1–2 vigorous sessions plus 3–4 moderate sessions per week<sup>46</sup>
<sup>^</sup>if known
<sup>#</sup> older heart age = heart age result is higher than chronological age

**Table 4:**
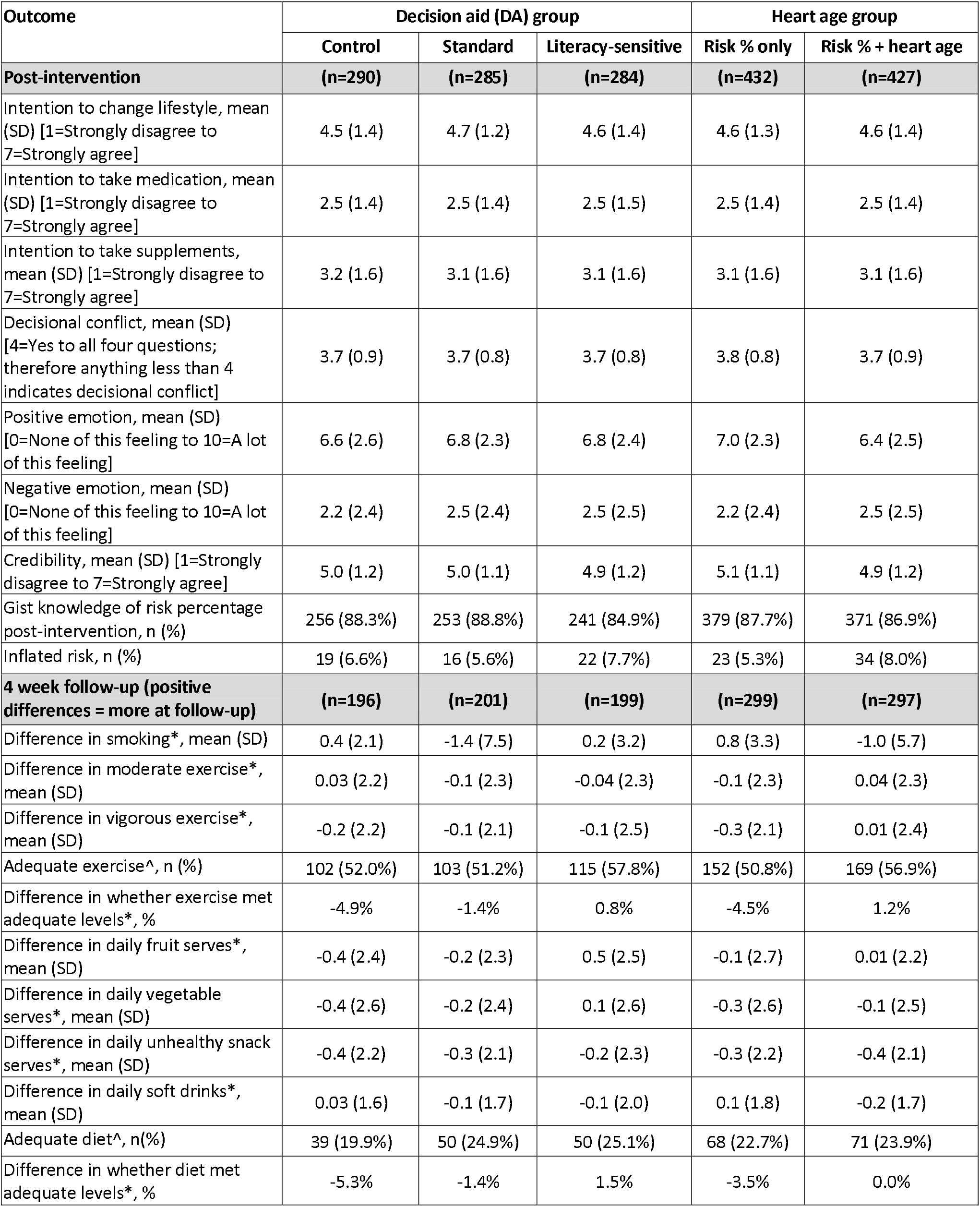
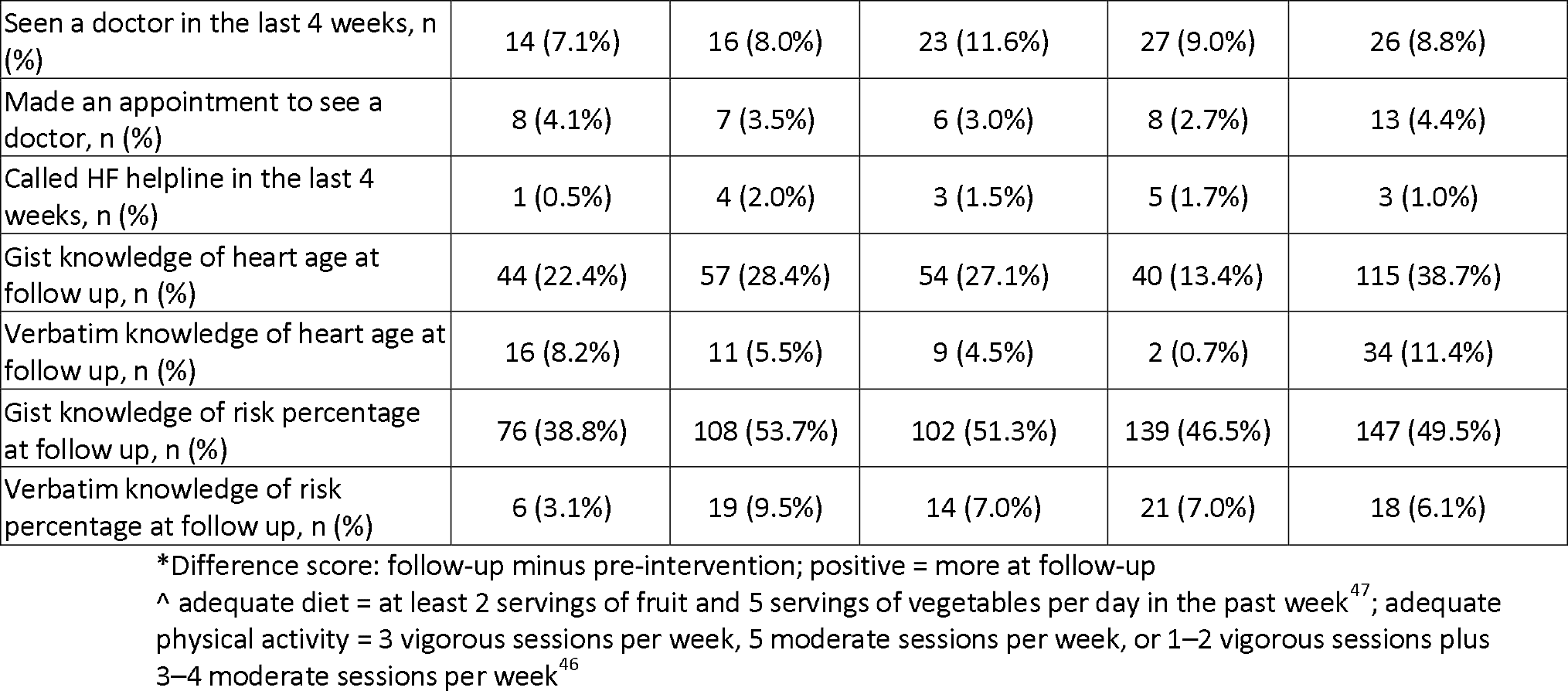
Trial outcomes by randomised group. Data are displayed as mean (standard deviation) unless otherwise indicated.

**Table 5:** Hypothesis 1: the decision aid groups will improve outcomes versus the control group.

|  | Outcome | Literacy-sensitive DA vs control<br>Mean difference (95% CI); p-value | Standard DA vs control<br>Mean difference (95% CI); p-value | Main effect<br>p-value |
| --- | --- | --- | --- | --- |
| Post-intervention | Intention to change lifestyle | 0.07 (-0.15, 0.29); p=.52 | 0.17 (-0.05, 0.39); p=.12 | .30 |
|  | Intention to talk to doctor about medication | 0.01 (-0.21, 0.24); p=.90 | 0.00 (-0.23, 0.22); p=.97 | .99 |
|  | Intention to take supplements | -0.10 (-0.36, 0.16); p=.43 | -0.09 (-0.34, 0.17); p=.52 | .70 |
|  | Decisional conflict + | 1.12 (0.72, 1.73); p=.62 | 0.98 (0.63, 1.54); p=.93 | .82 |
|  | Positive emotion | 0.16 (-0.23, 0.55); p=.43 | 0.31 (-0.08, 0.70); p=.12 | .29 |
|  | Negative emotion | 0.28 (-0.11, 0.68); p=.16 | 0.20 (-0.19, 0.60); p=.31 | .34 |
|  | Credibility | -0.12 (-0.30, 0.07); p=.22 | 0.01 (-0.18, 0.19); p=.95 | .34 |
|  | Gist knowledge of risk percentage post-intervention # | 1.22 (0.74, 2.02); p=.44 | 1.05 (0.63, 1.73); p=.85 | .72 |
|  | Inflated risk + | 1.10 (0.63, 1.92); p=.74 | 0.74 (0.39, 1.38); p=.34 | .42 |
| Follow-up (after 4 weeks, controlling for pre-intervention) | Daily smoking (no. cigarettes) * | 0.41 (-2.34, 3.16); p=.77 | -1.48 (-4.17, 1.20); p=.28 | .29 |
|  | Weekly vigorous exercise sessions * | 0.23 (-0.15, 0.62); p=.24 | 0.00 (-0.38, 0.38); p=.99 | .39 |
|  | Weekly moderate exercise sessions * | 0.03 (-0.36, 0.42); p=.89 | -0.03 (-0.42, 0.36); p=.87 | .95 |
|  | Whether exercise met adequate levels + | 1.10 (0.95, 1.28); p=.19 | 1.04 (0.89, 1.21); p=.64 | .41 |
|  | Daily fruit serves * | **0.69 (0.32, 1.06); p<.001** | 0.21 (-0.13, 0.55); p=.23 | **<.001** |
|  | Daily vegetable serves * | 0.38 (-0.03, 0.78); p=.07 | 0.04 (-0.36, 0.43); p=.85 | .13 |
|  | Daily unhealthy snack serves * | 0.11 (-0.17, 0.40); p=.43 | 0.02 (-0.26, 0.31); p=.87 | .71 |
|  | Daily soft drink serves * | 0.12 (-0.10, 0.35); p=.28 | 0.05 (-0.17, 0.27); p=.65 | .55 |
|  | Whether diet met adequate levels + | 1.23 (0.87, 1.74); p=.23 | 1.16 (0.83, 1.62); p=.37 | .48 |
| Follow-up only (after 4 weeks) | Has seen a doctor in the last 4 weeks+ | 1.60 (0.85, 3.02); p=.14 | 1.04 (0.52, 2.07); p=.92 | .23 |
|  | Intends to see a doctor at follow up + | 0.75 (0.27, 2.10); p=.58 | 0.86 (0.31, 2.41); p=.78 | .86 |
|  | Has called HF helpline in the last 4 weeks + | 3.00 (0.31, 29.07); p=.34 | 3.81 (0.45, 32.25); p=.22 | .47 |
|  | Gist knowledge of heart age at follow up + | 1.12 (0.80, 1.56); p=.51 | 1.16 (0.84, 1.61); p=.36 | .65 |
|  | Verbatim knowledge of heart age at follow up + | 0.47 (0.22, 1.03); p=.06 | 0.58 (0.28, 1.20); p=.14 | .12 |
|  | Gist knowledge of risk percentage at follow up + | **1.28 (1.04, 1.58); p=.021** | **1.41 (1.14, 1.74); p=.002** | **.006** |
|  | Verbatim knowledge of risk percentage at follow up + | 2.34 (0.91, 6.05); p=.08 | **3.25 (1.31, 8.07); p=.011** | **.039** |

+ analysis by modified Poisson regression, data shown as **incidence rate ratios**

**Figure 3:**
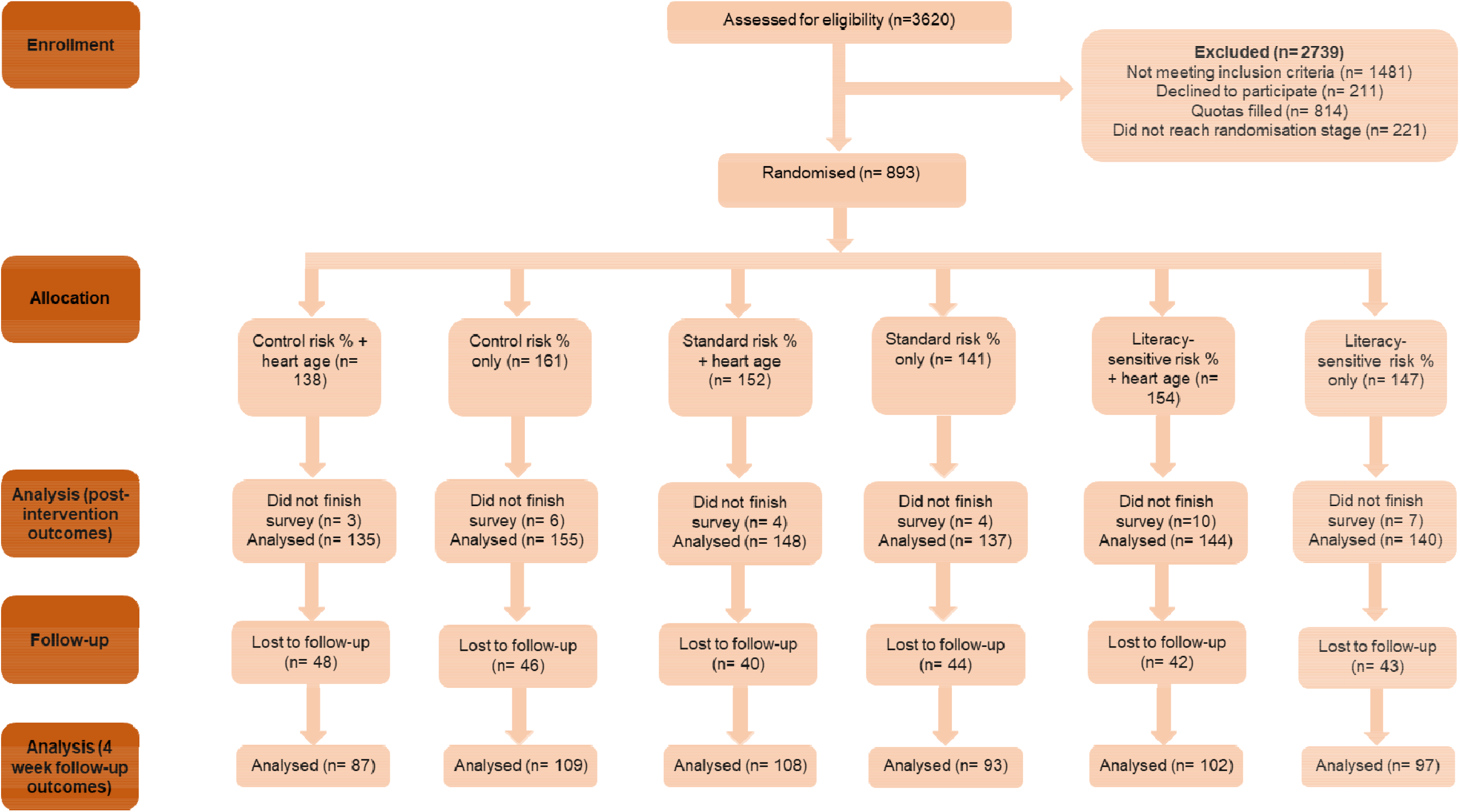
CONSORT diagram.

#### Post-intervention differences between decision aid groups

Immediately post-intervention, there were no differences between the 3 decision aid groups for the primary outcome of lifestyle intentions, or secondary outcomes of risk perception, credibility, emotional response or decisional conflict. For Hypothesis 1, the combined decision aid groups were no different to the control condition for any outcome (Table 5). For Hypothesis 2, there was no difference between the standard and literacy-sensitive decision aids for any outcome (Table 6). There were significant interactions between decision aid and health literacy for intention to talk to a doctor about medication (p=0.019) and emotional response (positive p=0.010; negative p=0.006). Participants with lower health literacy who received the literacy-sensitive decision aid had a more negative/less positive emotional response and had stronger intentions to see a doctor about medication, compared to the other groups (Table 6).

**Table 6:** Hypothesis 2: the literacy-sensitive decision aid will improve outcomes versus the standard decision aid regardless of health literacy level.

|  | Outcome | Standard DA (vs literacy-sensitive)<br>Estimated difference (95%CI); p-value | NVS_score x Group interaction<br>p-value |
| --- | --- | --- | --- |
| **Post-intervention)** | Intention to change lifestyle | 0.10 (-0.12, 0.32); p=.37 | .22 |
|  | Intention to talk to doctor about medication | -0.02 (-0.24, 0.21); p=.87 | **.019** |
|  | Intention to take supplements | 0.02 (-0.24, 0.28); p=.90 | .10 |
|  | Decisional conflict + | 0.88 (0.57, 1.36); p=.56 | .53 |
|  | Positive emotion | 0.16 (-0.24, 0.55); p=.44 | **.010** |
|  | Negative emotion | -0.08 (-0.48, 0.32); p=.69 | **.006** |
|  | Credibility | 0.12 (-0.06, 0.31); p=.20 | .11 |
|  | Gist knowledge of risk percentage post-intervention # | 0.86 (0.52, 1.41); p=.55 | **.007** |
|  | Inflated risk perception (above actual level) + | 0.70 (0.37, 1.23); p=.20 | .72 |
| **Follow-up (after 4 weeks, controlling for pre-intervention)** | Daily smoking (no. cigarettes) * | -1.90 (-4.33, 0.53); p=.13 | .90 |
|  | Weekly vigorous exercise sessions * | -0.23 (-0.62, 0.16); p=.24 | .20 |
|  | Weekly moderate exercise sessions * | -0.06 (-0.45, 0.32); p=.76 | .50 |
|  | Whether exercise met adequate levels + | 0.94 (0.81, 1.09); p=.43 | .35 |
|  | Daily fruit serves * | **-0.48 (-0.86, -0.11); p=.012** | .15 |
|  | Daily vegetable serves * | -0.34 (-0.74, 0.06); p=.10 | .10 |
|  | Daily unhealthy snack serves * | -0.09 (-0.38, 0.20); p=.53 | .97 |
|  | Daily soft drink serves * | -0.07 (-0.30, 0.16); p=.53 | .77 |
|  | Whether diet met adequate levels + | 0.94 (0.69, 1.28); p=.71 | .90 |
| **Follow-up (after 4 weeks)** | Has seen a doctor in the last 4 weeks+ | 0.65 (0.35, 1.19); p=.16 | .75 |
|  | Intends to see a doctor at follow up + | 1.15 (0.39, 3.36); p=.80 | Not tested (insufficient variability) |
|  | Has called HF helpline in the last 4 weeks+ | 1.27 (0.26, 6.09); p=.77 | **<.001** |
|  | Gist knowledge of heart age at follow up + | 1.04 (0.77, 1.41); p=.81 | .61 |
|  | Verbatim knowledge of heart age at follow up + | 1.24 (0.53, 2.89); p=.62 | .27 |
|  | Gist knowledge of risk percentage at follow up + | 1.10 (0.92, 1.30); p=.29 | .83 |
|  | Verbatim knowledge of risk percentage at follow up + | 1.39 (0.71, 2.69); p=.33 | **<.001** |

+ analysis by modified Poisson regression, data shown as **incidence rate ratios**

#### 4 week differences between decision aid groups

At follow-up after 4 weeks, there were no significant differences between the control and decision aid groups for most self-reported behaviours. However, the literacy-sensitive decision aid group had higher fruit consumption compared to both control (difference in predicted counts = 0.69 (95%CI: 0.32 to 1.06); p<.001) and the standard decision aid group (difference in predicted counts = 0.48 (95%CI: 0.11 to 0.86); p=.012). The decision aid groups were both more likely to know whether their risk was low, medium or high than the control group (literacy-sensitive DA: IRR=1.28 (95%CI: 1.04 to 1.58); p=.021; standard DA: IRR=1.41 (95%CI: 1.14 to 1.74); p=.002). The standard DA group was more likely to know their exact risk percentage result compared to control (IRR=3.25 (95%CI: 1.31 to 8.07); p=.011); see Table 5. There were significant differences between decision aid groups by health literacy levels for self-reported calls to the Heart Foundation helpline (p<0.001) and verbatim knowledge of CVD percentage risk at follow up (p<0.001). No one with low health literacy reported calling the helpline or remembered their exact CVD risk in the control group. The standard decision aid increased both outcomes in all health literacy groups, and the literacy-sensitive decision aid increased both outcomes in low and high health literacy groups but not medium (Table 6).

#### Post-intervention differences between heart age groups

Immediately post-intervention, there were no differences between the two heart age groups for the primary outcome of lifestyle intentions, or secondary outcomes of risk perception or decisional conflict. For Hypothesis 3, the heart age group was less likely to have a positive emotional response (mean difference = -0.56 (95%CI: -0.88, -0.24); p=.001), less likely to perceive the message as credible (−0.20 (−0.35, -0.05); p=.010), and more likely to know whether their risk was low, medium or high (2.03 (1.33, 3.08); p=.001), compared to the percentage risk only group (Table 7). When the heart age result was older, there were significant differences indicating less positive (−0.75 (−1.19, - 0.31); p=.001) and more negative (0.57 (0.12, 1.02); p=.014) emotional responses, lower credibility (−0.29 (−0.49, -0.09); p=.005) and higher perceived risk level (2.11 (1.31, 3.39); p=.002) when heart age was shown. No such differences were found for those who received the same age or younger results (Table 7).

**Table 7:** Hypothesis 3: adding heart age to percentage risk will improve outcomes versus percentage risk only.

|  | Outcome | Heart age shown vs not shown |  |  |
| --- | --- | --- | --- | --- |
|  |  | Across all participants | Older heart age result | Same/younger heart age result |
| Post-intervention) | Intention to change lifestyle | -0.04 (-0.22, 0.14); p=.64 | -0.11 (-0.34, 0.13); p=.36 | 0.02 (-0.25, 0.30); p=.87 |
|  | Intention to take medication | .04 (-0.15, 0.22); p=.70 | 0.11 (-0.13, 0.36); p=.37 | -0.14 (-0.41, 0.13); p=.31 |
|  | Intention to take supplements | 0.00 (-0.21, 0.21); p=.99 | 0.07 (-0.21, 0.35); p=.63 | -0.14 (-0.47, 0.19); p=.41 |
|  | Decisional conflict + (%) | 1.27 (0.89, 1.83); p=.19 | 1.08 (0.72, 1.62); p=.71 | 1.72 (0.81, 3.67); p=.16 |
|  | Positive emotion | **-0.56 (-0.88, -0.24); p=.001** | **-0.75 (-1.19, -0.31); p=.001** | -0.25 (-0.70, 0.20); p=.28 |
|  | Negative emotion | 0.26 (-0.06, 0.58); p=.12 | **0.57 (0.12, 1.02); p=.014** | -0.27 (-0.71, 0.17); p=.23 |
|  | Credibility | **-0.20 (-0.35, -0.05); p=.010** | **-0.29 (-0.49, -0.09); p=.005** | -0.06 (-0.29, 0.17); p=.60 |
|  | Gist knowledge of risk percentage post-intervention # | **2.03 (1.33, 3.08); p=.001** | **2.12 (1.32, 3.41); p=.002** | 1.60 (0.58, 4.37); p=.36 |
|  | Inflated risk + (%) | 1.60 (0.98, 2.61); p=.058 | 1.70 (0.93, 3.13); p=.086 | 1.38 (0.57, 3.36); p=.47 |
| Follow-up (after 4 weeks, controlling for pre-intervention) | Daily smoking (no. cigarettes) * | -0.77 (-2.93, 1.40); p=.49 | Not estimated (n=4 not shown) | -0.66 (-2.94, 1.61); p=.57 |
|  | Weekly vigorous exercise sessions * | 0.29 (-0.02, 0.60); p=.07 | **0.58 (0.09, 1.07); p=.021** | 0.04 (-0.37, 0.44); p=.85 |
|  | Weekly moderate exercise sessions * | 0.05 (-0.26, 0.37); p=.74 | 0.45 (-0.01, 0.91); p=.056 | -0.26 (-0.72, 0.20); p=.27 |
|  | Whether exercise met adequate levels + | 1.16 (0.99, 1.26); p=.08 | **1.23 (1.05, 1.45); p=.010** | 1.03 (0.86, 1.24); p=.74 |
|  | Daily fruit serves * | 0.02 (-0.28, 0.31); p=.92 | 0.42 (-0.06, 0.89); p=.084 | -0.26 (-0.63, 0.11); p=.17 |
|  | Daily vegetable serves * | 0.30 (-0.02, 0.63); p=.07 | **0.57 (0.05, 1.09); p=.032** | -0.14 (-0.28, 0.56); p=.51 |
|  | Daily unhealthy snack serves * | -0.05 (-0.28, 0.18); p=.68 | 0.22 (-0.15, 0.58); p=.25 | -0.28 (-0.58, 0.02); p=.067 |
|  | Daily soft drink serves * | -0.14 (-0.33, 0.04); p=.13 | 0.03 (-0.22, 0.27); p=.83 | **-0.34 (-0.61, -0.07); p=.012** |
|  | Whether diet met adequate levels + | 1.14 (0.87, 1.50); p=.34 | **1.48 (1.00, 2.18); p=.048** | 0.95 (0.66, 1.38); p=.79 |
| Follow-up (after 4 weeks) | Has seen a doctor in the last 4 weeks+ | 0.99 (0.60, 1.63); p=.96 | 0.81 (0.37, 1.80); p=.61 | 1.15 (0.58, 2.26); p=.69 |
|  | Intends to see a doctor at follow up + | 1.61 (0.67, 3.84); p=.29 | 0.67 (0.17, 0.27); p=.58 | 4.17 (0.90, 19.32); p=.068 |
|  | Has called HF helpline in the last 4 weeks+ | 0.65 (0.17, 2.53); p=.54 | 1.23 (0.25, 6.03); p=.80 | **2.66 (1.76, 4.03); p<.001** |
|  | Gist knowledge of heart age at follow up + | **2.90 (2.10, 3.99); p<.001** | **3.38 (2.05, 5.55); p<.001** | **6.67 (1.50, 32.41); p=.013** |
|  | Verbatim knowledge of heart age at follow up + | **18.13 (4.36, 75.48); p<.001** | Not estimated (n=2 not shown) | Not estimated (n=2 not shown) |
|  | Gist knowledge of risk percentage at follow up + | 1.11 (0.95, 1.30); p=.20 | 1.09 (0.91, 1.29); p=.35 | 1.16 (0.87, 1.55); p=.31 |
|  | Verbatim knowledge of risk percentage at follow up + | 0.82 (0.44, 1.50); p=.52 | 1.02 (0.40, 2.57); p=.97 | 0.68 (0.31, 1.52); p=.35 |

+ analysis by modified Poisson regression, data shown as **incidence rate ratios**; # analysis by ordered logistic regression, data shown as odds ratio of being in next highest (odds in heart age, relative to not shown); \* analysis by negative binomial regression, data shown as **differences in predicted counts**; & Unstable estimate: 5/299 individuals not shown heart age used helpline, compared to 3/297 shown heart age.

#### 4 week differences between heart age groups

At 4 week follow-up there were no significant differences between the heart age groups in terms of lifestyle behaviour change, seeing a doctor for a Heart Health Check or gist knowledge of risk level (Table 7). Unsurprisingly, being shown heart age led to greater gist knowledge of heart age (2.90 (2.10, 3.99); p<.001) and verbatim knowledge of heart age (18.13 (4.36, 75.48); p<.001) compared to the those who weren’t shown their heart age, but there was no difference between the heart age and percentage risk only groups for knowledge of percentage risk. Within the heart age group who saw both risk formats, participants were more likely to have verbatim knowledge of their heart age (11%) than their percentage risk (6%) (McNemar’s chi-squared test for paired proportions: χ^2 (1)=6.10, p=.014, difference in proportions: 5.4%, 95%CI: 0.8% to 10.0%). When the heart age result was older, there were significant differences indicating more vigorous exercise (0.58 (0.09, 1.07); p=.021), more vegetable serves (0.57 (0.05, 1.09); p=.032), higher chance of meeting guidelines for exercise (1.23 (1.05, 1.45); p=.010) and diet (1.48 (1.00, 2.18); p=.048), when heart age was shown. When the heart age result was the same or younger than current age, there were significant differences indicating fewer soft drink serves (−0.34 (−0.61, -0.07); p=.012), and higher chance of calling the Heart Foundation helpline (12.66 (1.76, 4.03); p<.001), when heart age was shown (Table 7).

### Stage 3

As part of the follow up survey, participants were asked if they’d like to be interviewed about the risk calculator. Twenty-seven of those indicated that they would. From this pool, 20 were selected to represent a range of ages, genders, risk levels, and health literacy. The interviews were done in four stages so that any user feedback from the interviews could be discussed amongst the team (CB, CAB and JA) and then implemented into the calculator for the next interviews, in an iterative process. The issues addressed in each round of interviews are shown in Appendix 2.

## DISCUSSION

We used both a mixed methods development and evaluation process to produce a CVD decision aid that is effective for improving verbatim and gist knowledge of CVD risk and fruit consumption after 4 weeks. The resulting intervention is a scalable ehealth tool suitable for people with varying health literacy. This consumer tool will supplement a GP version to use within consultations^36,37^, providing GPs with a clear action for their patients to follow up when lifestyle change is recommended.

The paper provides an example of how to apply literacy-sensitive design principles to evidence-based decision making and behaviour change tools. The results show literacy-sensitive decision aids can support people with lower health literacy to make informed decisions, while still being suitable for the general population. A recent review of decision aids for people with lower health literacy^53,54^ showed that decision aids that used health literacy design strategies led to improved knowledge, decisional conflict and decision making outcomes. Further, decision aids that used explicit strategies to reduce cognitive burden showed greater improvements knowledge for people with low health literacy and from disadvantaged backgrounds^54^. The review highlighted the need for more consideration of health literacy in decision aid development. This study addresses the findings in the context of CVD prevention for the first time.

We observed a number of interactions with health literacy, showing the importance of considering this as a covariate when investigating shared decision making and behaviour change outcomes. The literacy-sensitive version of the decision aid produced more negative emotional responses and greater intentions to speak to a doctor about medication options to reduce CVD risk, amongst those with lower health literacy. This may reflect risk and choice awareness in this group, if they had not previously considered themselves to have risk factors for heart disease that could be addressed with preventive medication. As this sample was predominantly low risk, we would not want a decision aid to lead to greater actual medication uptake in this group, but speaking with a doctor about risk and how to reduce it may be a positive outcome in line with guidelines to assess risk in this age group^1^. We replicated previous decision aid studies in finding increased knowledge of risk amongst the decision aid groups compared to control^35^. We also replicated our previous finding that a literacy-sensitive action plan can improve diet outcomes across different levels of health literacy, although this was more marked for people with low health literacy^29,55^.

This study also replicated a number of heart age effects found in reviews of previous research, in that it leads to a more negative emotional response, increased gist and verbatim knowledge of heart age but not percentage risk, and reduced credibility, but is neutral for lifestyle change overall^56,57^. Our subgroup analyses suggest that more nuanced study designs may be required to better understand the effects of heart age. Firstly, amongst those who were shown their heart age, gist knowledge of percentage risk was initially improved, but after 4 weeks gist and verbatim knowledge were higher for heart age than for percentage risk. Previous studies have shown that people receiving an older heart age may react defensively and focus on other information, such as a low short-term risk level, which in turn may reduce credibility of the risk result^24,58^. Analyses of people who received an older heart age result suggests it may be useful as a marketing tool to get attention and initiate behaviour change, but knowledge of heart age did not translate to knowledge of risk. For the intended purpose of a decision aid to be used in a clinical context, the focus needs to be on validated risk results to make informed decisions about medication. For this reason, we made the decision to use the non-heart age version of the literacy-sensitive decision aid for future research in general practice. However, online heart age tools could incorporate decision aid and action plan elements with no detrimental effects. Future trials need to be designed differently to isolate older heart age results and follow-up behaviour over time. In considering how to power such trials, researchers will need to consider how the specific heart age tool they are using is calibrated for the intended population (e.g. ∼50% older in our sample using NZ method versus ∼80% in Australian/UK HF tool^23,24^). The primary outcome also needs to carefully considered. Most heart age research has been done with a primary outcome of immediate lifestyle change intentions, where we found no differences. More research could be done to verify the self-reported behaviour change amongst people receiving older heart age results we observed after 4 weeks, using more objective measures such as pedometers.

The end user interviews were helpful for improving simple navigation and wording issues in the literacy-sensitive version of the decision aid, but there were some larger issues that cannot be resolved in an online tool. Most users did not know their blood pressure or cholesterol results, but even if they had been assessed recently, they had difficulty understanding where the different numbers should be entered. This was particularly difficult for cholesterol results in pathology test reports. We will therefore test the final revised tool in clinical practice to address the issue of unknown blood pressure and cholesterol which reduces the accuracy, and limit display of options in line with current medication guidelines.

## Strengths and limitations

A major strength of this study is that we were able to recruit a large, diverse sample in terms of health literacy and risk results. We had sufficient follow-up to run the study per protocol despite COVID-19 disruptions, and observed no difference in dropouts for key variables. A limitation is that the online panel sample may not be representative of the general population, and may better reflect users of online heart age tools than patients presenting to primary care for CVD risk assessment. Different countries use also different CVD risk models/heart age algorithms, which may affect the results given the differences we observed in the older heart age sample. Finally, we used validated outcomes where possible, but behaviour change was self-reported. Future research on heart age could consider using objective measures over time.

## Conclusion

This study shows the value of combining health literacy-sensitive design with best practice risk communication and behaviour change tools. Although aimed at addressing the needs of people with lower health literacy, this approach improved knowledge of CVD risk and heart age, and behaviour, in a sample with varying health literacy levels. The role of heart age remains somewhat unclear, with both advantages and disadvantages, but no clear evidence of an effect on lifestyle change intentions or behaviour overall. Further research should investigate implementation pathways for integrating such consumer tools with clinical practice, and distinguish between older and younger heart age results.

## Data Availability

Presently the data is stored securely at the University of Sydney's server. Should you wish to ask about accessing any data, please contact Dr Carissa Bonner at

## Acknowledgments

This study was funded by a Vanguard Grant from the National Heart Foundation of Australia (ID 102215)

## Notes

### Competing Interest Statement

The authors have declared no competing interest.

### Clinical Trial

ACTRN12620000806965

### Funding Statement

This was work was conducted as a result of Dr Carissa Bonner's Vanguard Grant from the National Heart Foundation of Australia

### Author Declarations

University of Sydney Human Research Ethics Committee

